# Youth Psychotic Experiences: Diagnostic Associations and Evaluation of the CAPE-16

**DOI:** 10.1101/2024.04.18.24306017

**Authors:** Viktoria Birkenæs, Pravesh Parekh, Laura Hegemann, Nora R. Bakken, Evgeniia Frei, Piotr Jaholkowski, Olav B. Smeland, Ezra Susser, Katrina M. Rodriguez, Markos Tesfaye, Ole A. Andreassen, Alexandra Havdahl, Ida E. Sønderby

**Author notes:** **Correspondence to** Viktoria Birkenæs, Centre for Precision Psychiatry, Division of Mental Health and Addiction, Oslo University Hospital, Ullevål, Kirkeveien 166, Building 49, 0450 Oslo, Norway. Contributed equally to this work (Alexandra Havdahl, Ida E. Sønderby).

## Abstract

**Background:** Adolescent self-reported psychotic experiences are associated with mental illness and could help guide prevention strategies. The Community Assessment of Psychic Experiences (CAPE) was developed over 20 years ago. In a rapidly changing society, where new generations of adolescents are growing up in an increasingly digital world, it is crucial to ensure high reliability and validity of the questionnaire.

**Methods:** In this observational validation study, we used unique transgenerational questionnaire and health registry data from the Norwegian Mother, Father, and Child Cohort, a population-based pregnancy cohort. Adolescents, aged ∼14 years, responded to the CAPE-16 (*n* = 18,835) and fathers to the CAPE-9 questionnaire (*n* = 28,793). We investigated the psychometric properties of CAPE-16 through factor analyses, measurement invariance testing across biological sex, response before/during the COVID-19 pandemic, and generations (comparison with fathers), and examined associations with later psychiatric diagnoses.

**Outcomes:** One third (33·4%) of adolescents reported lifetime psychotic experiences. We confirmed a three-factor structure (paranoia, bizarre thoughts, and hallucinations) of CAPE-16, and observed good scale reliability of the distress and frequency subscales (ω = ·86 and ·90). CAPE-16 measured psychotic experiences were invariant to biological sex and pandemic status. CAPE-9 was non-invariant across generations, with items related to understanding of the digital world (*electrical influences*) prone to bias. CAPE-16 sum scores were associated with a subsequent psychiatric diagnosis, particularly psychotic disorders (*frequency:* OR = 2·06; 97·5% CI = 1·70–2·46; *distress:* OR = 1·93; 97·5% CI = 1·63–2·26).

**Interpretation:** CAPE-16 showed robust psychometric properties across sex and pandemic status, and sum scores were associated with subsequent psychiatric diagnoses, particularly psychotic disorders. These findings suggest that with certain adjustments, CAPE-16 could have value as a screening tool for adolescents in the modern, digital world.

**Funding:** European Union’s Horizon 2020 Programme, Research Council of Norway, South-Eastern Norway Regional Health Authority, NIMH, and the KG Jebsen Stiftelsen.

## Introduction

Psychotic experiences (PEs) are delusions and hallucinations that do not reach the clinical threshold for a psychotic episode.^1^ The estimated prevalence of PEs is around 8-17% in children and adolescents^2^ compared to 5-10% in adults.^1,3^ In adults, PEs are associated with both co-occurring and subsequent mental illness^4^. Similarly, in youth, PEs are associated with a four-fold increased risk of developing psychosis and a three-fold increased risk of non-psychotic mental illnesses.^2^ These findings highlight adolescent PEs as targets for early detection and intervention. Yet, to identify clinically relevant PEs, we need valid and reliable PE questionnaires.

One of the most used measures of PEs in general population samples is the Community Assessment of Psychic Experiences (CAPE),^5^ and particularly brief screening versions of the tool, such as CAPE-15. Evidence suggest that CAPE-15 best fits a three-factor structure consisting of persecutory ideation, bizarre experiences, and perceptual abnormalities.^6^ We previously verified this structure in a population-based sample of Norwegian fathers from the MoBa cohort using data from the self-reported nine-item CAPE (CAPE-9).^7^ Recently collected data from MoBa children now give us access to unique transgenerational measures of PEs. Yet, CAPE was developed more than 20 years ago.^5^ During the past two decades, Western societies have changed rapidly, and self-report tools like the CAPE may no longer accurately capture the psychotic experiences of modern adolescents. Because of the digital revolution and the introduction of the internet, smartphones, and social media, adolescents today inhabit a world different to the one their parents grew up in.^8^ These contextual disparities could potentially impact how youths perceive and respond to questions about their experiences. Therefore, it is crucial to ensure that the measurement tool being used is still valid and reliable.

Important difference within sub-groups of youths may also exist. During the past decade, there has been an apparent rise in affective symptoms among adolescents, particularly girls^9^ – a problem further exacerbated by the COVID-19 pandemic. The pandemic itself was likely an important stressor for those who experienced it, particularly adolescents.^10^ Yet, it is unknown if the pandemic has impacted the rate of PEs. Further, adolescent boys have an increased vulnerability for developing psychosis^11^ but studies examining sex differences in reported PEs yield inconsistent findings.^12^ Variability in CAPE-16’s measurement properties across sex, pandemic status, and generations could significantly affect the accuracy and consistency of self-reports across individuals and challenge interpretations of findings and the usefulness of the tool.

Questionnaire variability can be assessed through measurement invariance testing, which evaluates whether group differences are solely due to differences in scaling (i.e., one group simply reports more symptoms) or due to differences in the *pattern* of responses.^13^ Preliminary evidence suggests that CAPE measures PEs similarly across boys and girls^14^ but there is overall limited knowledge about potential sex differences, and we do not know if CAPE responses patterns are equivalent before and during the COVID-19 pandemic or across generations that grew up either before or in a “digital world”. This method may help us identify ways to improve the measure for future screening and prediction purposes in current youth populations.

Children reporting PEs have an increased likelihood of adult-onset psychotic disorders, as well as non-psychotic mental illness^2^. Still, few prospective studies have explored associations between adolescent PEs and subsequent mental illness.^2^ Moreover, the diagnostic associations of item-level PEs are largely unexplored.^3^ Characterizing the associations between specific PEs and psychiatric diagnoses may give insight into early manifestations of illness and help improve identification of those affected.

Here, we assessed PEs as measured by CAPE-16 during 2016-2022 in a prospective population-based sample of Norwegian adolescents. By validating CAPE-16, we aimed to determine its effectiveness in capturing experiences related to the psychosis continuum in different contexts and to inform research on prediction and early identification of psychosis. We first examined the psychometric properties of CAPE-16 by: (1) investigating factor structure and scale reliability, (2) assessing measurement invariance across biological sex, response before/during COVID-19, and generations (adolescents compared to fathers), and (3) assessing the associations between CAPE-16 and later psychiatric diagnoses, with a focus on psychotic disorders.

## Methods

### Participants

MoBa is a population-based pregnancy cohort study conducted by the Norwegian Institute of Public Health. Participants were recruited from all over Norway from 1999-2008. Mothers consented to participation in 41% of invited pregnancies. Fathers were first invited to contribute at week 15 of the pregnancy. ^15^ All participants provided informed written consent. The formation of MoBa and initial data collection were based on a license from the Norwegian Data Protection Agency and approval from the Regional Committees for Medical and Health Research Ethics. The Norwegian Health Registry Act regulates the MoBa cohort, and the Regional Committees for Medical and Health Research Ethics approved the current study (2016/1226/REK sør-øst C).

### Measures

#### Sociodemographic variables

Information about sex assigned at birth, birth year and weight, parity, and parent’s age at birth was collected from the Norwegian Medical Birth Registry, a national registry containing information about all births in Norway. Parents reported income and educational attainment in the baseline MoBa questionnaires.

#### Symptom measures

Between 2016 and 2022, all MoBa children (around 14 years at sampling) received health and lifestyle questionnaires. Approximately ∼23% (*n* = 21,122) responded to the questionnaires, including the CAPE-16 (complete cases: *n* = 18,835, details in Supplementary table S4). CAPE-16 comprises CAPE-15^16^ and an additional item (*delusions of reference*) from CAPE-9^7^ (overview of items in Supplementary Table S3). Each item is rated on a 4-point scale for both frequency of experiences and related distress (“never”, “sometimes”, “often”, “nearly always”). We defined our PE cut-off as scoring “often” or “nearly always” on one or more questions in accordance with previous literature^17^. During 2015, MoBa fathers responded to CAPE-9 (complete cases *n* = 28,793).^7^ When comparing fathers and adolescents, we selected the items comprising CAPE-9 from CAPE-16.

#### Emotional symptoms

Emotional symptoms, like anxiety and depression, typically accompany PEs^18^. Emotional symptoms affect the measurement validity of screening tools for other psychiatric conditions and experiences ^19^, and could potentially lead to inflated associations between PEs and mental diagnoses. We currently aim to assess the effectiveness of CAPE-16 in identifying experiences on the psychosis continuum beyond emotional symptoms. Hence, we included the Symptoms Checklist-10 (SCL-10)^20^ as a covariate.

#### Psychiatric diagnoses

Psychiatric diagnoses received after questionnaire response were derived from the Norwegian Patient Registry, a specialist health care registry containing ICD-10^21^ codes. Diagnoses included in the analyses were psychotic disorders (F20, F22, F23, F25, F28, F29), depressive episodes (F32), bipolar disorder (F31), phobias (F40), anxiety disorders (F40), obsessive-compulsive disorder (OCD; F42), trauma- and stressor-related disorders (F43.0, F43.1, F43.2), somatoform disorder (F45), eating disorders (F50), and personality disorders (F60). We included only unrelated individuals (details in Supplementary Figure S1).

### Statistical analyses

All analyses were performed in R, version 4.0.3^22^.

#### Scale reliability and confirmatory factor analysis

Scale reliability of the CAPE-16 subscales was evaluated using omega estimates^23^. We conducted confirmatory factor analyses (CFA) based on the previously observed three-factor structure of CAPE-15^6^ and CAPE-9^7^. To better handle violations of normality, we used a robust maximum likelihood estimator^24^.

#### Measurement invariance testing

We tested measurement invariance (MI) across group status (boys/girls, response before/during COVID-19, adolescents/fathers)^13^. This method involves testing models with increasingly strict parameter constraints to evaluate if parameters are equal across groups. We first tested the replicability of the factor structure (configural model), then constrained factor loadings to be equal across groups (metric invariance), and lastly constrained both intercepts and loadings (scalar invariance).

We evaluated the configural model based on overall model fit, with a good fit indicated by comparative fit index (CFI) and Tucker Lewis index (TLI) >0·95, root mean square error of approximation (RMSEA) <0·06, and standardized root mean square residual (SRMR) <0·08. To evaluate metric and scalar invariance, we compared models by estimating change in model fit. Due to the large sample size, we used a stricter cut-off ≥-·01 for ΔCFI and ΔTFI, ≥·015 in ΔRMSEA and ≥·030 in ΔSRMR for metric invariance, and ≥·015 in ΔSRMR for scalar invariance^13^.

We further assessed partial MI. This involves trying to establish invariance among a subset of specified parameters if the complete model is not established. The model parameters chosen to test possible partial invariance were based on modification indices (i.e., the amount the chi-square fit would reduce if the constraint on that parameter were removed) ^13^. Unequal samples may result in less sensitive MI tests^25^. Hence, as a sensitivity test, we randomly drew fathers to create a sample matching the number of adolescents.

#### CAPE-16 and subsequent psychiatric diagnoses

We performed binominal logistic regression to test whether adolescents’ CAPE-16 sum scores and single items were associated with later psychiatric diagnoses. Frequency and distress sum scores were standardized with *Z*-transformation. Second, we included current emotional symptoms (SCL-10 sum scores) as a covariate. *P*-values were adjusted for multiple comparisons using Benjamini-Hochberg correction^26^. Approximately 23% of the initial MoBa children participated in the current wave.

Therefore, to evaluate potential attrition bias, we re-ran our logistic analyses with inverse probability weights (IPW; described in Supplementary Note 1).

#### Predicting psychotic disorders using CAPE-16

We used a machine learning approach to test if CAPE-16 could predict psychotic disorders (details in Supplementary Note 2). This explorative analysis was performed to investigate the validity of CAPE-16. If CAPE-16 successfully predicts diagnosed psychosis above chance levels, this would add evidence supporting its ability to measure psychosis-related phenomena. We included 49 features: all frequency and distress items, interactions between corresponding scale items, and biological sex. We used the RUSBoost algorithm, which is designed to handle imbalanced sample sizes between outcome categories. Overall, we ran four models: two with the complete sample and two where non-psychotic psychiatric diagnoses were removed (*n* = 14,852) – one of each model including regression of current emotional symptoms. We conducted 100 permutation tests to see if prediction performances were above chance level.

## Results

### Sample characteristics

Of the adolescents, 53·7% were registered as female in the birth registry. Mean age at response was 14·4 years (±·51). Fourteen percent reported any lifetime nicotine use, 17% alcohol, 1·4% cannabis, and 1·3% other drug use. Respondents and their parents had fewer psychiatric diagnoses than non-respondents, and parents of respondents were older and more highly educated than parents of non-respondents (Supplementary Tables S1-S2). In the fathers, mean age at response was 42.4 (±5·6). ^7^

#### Psychotic experiences in a Norwegian adolescent sample

One third (33·4%) of adolescents reported experiencing one or more PEs “often” or “nearly always” (Figure 1; Supplementary Table S4). Girls reported significantly more frequent (*r* = ·26, *p*<·0001) and distressing (*r* = ·27, *p<·0001*) PEs. There were significant, but small, positive associations between CAPE-16 sum scores and later year of birth (frequency: *r* = ·05, *p*<·0001; distress: *r* = 0·04, *p*<·0001) and between CAPE-16 and responding during the COVID-19 pandemic (frequency: *r* = ·05, *p*<·0001; distress: *r* = ·04, *p*<·0001).

**Figure 1.**
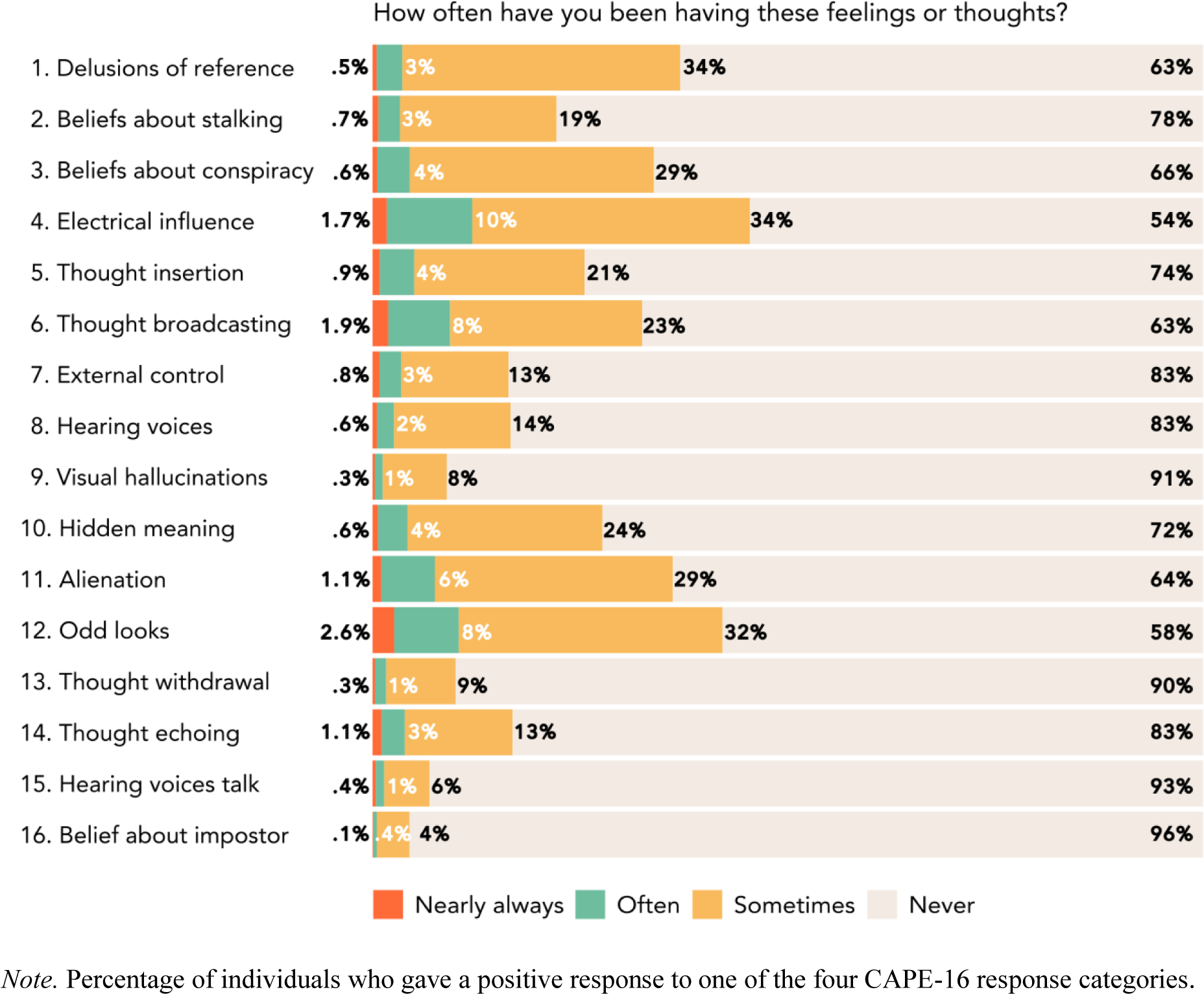
CAPE-16 frequency response patterns in MoBa adolescents

### Scale reliability and confirmatory factor analysis

Omega estimates were ·86 and ·90 for the frequency and distress subscales, respectively, suggesting excellent scale reliability^27^.

Table 1 provides CFA estimates for all subgroups. The CAPE-16 frequency and distress scales showed a good fit with the proposed three-factor structure^6^; persecutory ideation, bizarre thoughts, and hallucinations, with significant loadings for all items. The factors were replicated across biological sex, pandemic status, and generations.

**Table 1.**
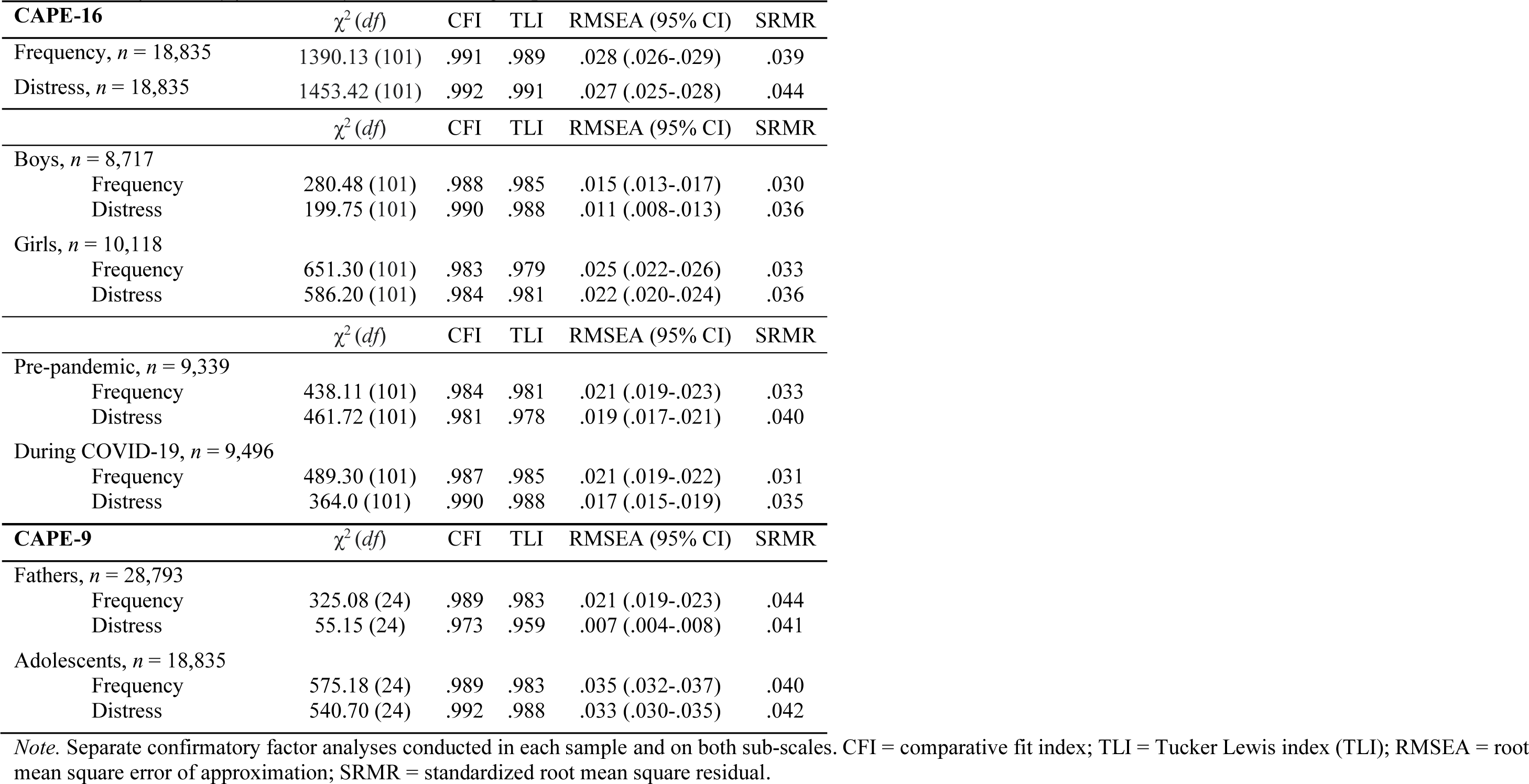
Confirmatory factor analyses in all sub-groups.

| **CAPE-16** | $\chi^2 (df)$ | CFI | TLI | RMSEA (95% CI) | SRMR |
| --- | --- | --- | --- | --- | --- |
| Frequency, $n = 18,835$ | 1390.13 (101) | .991 | .989 | .028 (.026-.029) | .039 |
| Distress, $n = 18,835$ | 1453.42 (101) | .992 | .991 | .027 (.025-.028) | .044 |
| | $\chi^2 (df)$ | CFI | TLI | RMSEA (95% CI) | SRMR |
| Boys, $n = 8,717$ | | | | | |
| Frequency | 280.48 (101) | .988 | .985 | .015 (.013-.017) | .030 |
| Distress | 199.75 (101) | .990 | .988 | .011 (.008-.013) | .036 |
| Girls, $n = 10,118$ | | | | | |
| Frequency | 651.30 (101) | .983 | .979 | .025 (.022-.026) | .033 |
| Distress | 586.20 (101) | .984 | .981 | .022 (.020-.024) | .036 |
| | $\chi^2 (df)$ | CFI | TLI | RMSEA (95% CI) | SRMR |
| Pre-pandemic, $n = 9,339$ | | | | | |
| Frequency | 438.11 (101) | .984 | .981 | .021 (.019-.023) | .033 |
| Distress | 461.72 (101) | .981 | .978 | .019 (.017-.021) | .040 |
| During COVID-19, $n = 9,496$ | | | | | |
| Frequency | 489.30 (101) | .987 | .985 | .021 (.019-.022) | .031 |
| Distress | 364.0 (101) | .990 | .988 | .017 (.015-.019) | .035 |
| **CAPE-9** | $\chi^2 (df)$ | CFI | TLI | RMSEA (95% CI) | SRMR |
| Fathers, $n = 28,793$ | | | | | |
| Frequency | 325.08 (24) | .989 | .983 | .021 (.019-.023) | .044 |
| Distress | 55.15 (24) | .973 | .959 | .007 (.004-.008) | .041 |
| Adolescents, $n = 18,835$ | | | | | |
| Frequency | 575.18 (24) | .989 | .983 | .035 (.032-.037) | .040 |
| Distress | 540.70 (24) | .992 | .988 | .033 (.030-.035) | .042 |

*Note.* Separate confirmatory factor analyses conducted in each sample and on both sub-scales. CFI = comparative fit index; TLI = Tucker Lewis index (TLI); RMSEA = root mean square error of approximation; SRMR = standardized root mean square residual.

### Measurement invariance across biological sex, pandemic status, and generations

We found evidence for full invariance across biological sex and pandemic status for both subscales, indicating that responses are comparable across groups (Tables 2a and 2b).

**Table 2a.** Frequency sub-scale: Test of measurement invariance across biological sex, pandemic status, and generations.

| **A. CAPE-16: Boys v. girls** |  |  |  |  |  |  |  |  |  |  |  |
| --- | --- | --- | --- | --- | --- | --- | --- | --- | --- | --- | --- |
| Model | $\chi^2 (df)$ | CFI | TLI | RMSEA (95% CI) | SRMR | $\Delta\chi^2 (\Delta df)$ | $\Delta CFI$ | $\Delta TLI$ | $\Delta RMSEA$ | $\Delta SRMR$ | Decision |
| M1: Configural Invariance | 931.78(202) | .984 | .981 | .021 (.019-.022) | .029 | — | — | — | — | — | — |
| M2: Metric Invariance | 1087.59(215)** | .981 | .979 | .022 (.021-.023) | .032 | 155.81 (13)** | -.003 | -.002 | .001 | .003 | Accept |
| M3: Scalar Invariance | 1427.05 (218)** | .974 | .973 | .025 (.024-.026) | .035 | 339.46 (13)** | -.007 | -.006 | .003 | .003 | Accept |
| **B. CAPE-16: Pre- v. during COVID-19** |  |  |  |  |  |  |  |  |  |  |  |
| Model | $\chi^2 (df)$ | CFI | TLI | RMSEA (95% CI) | SRMR | $\Delta\chi^2 (\Delta df)$ | $\Delta CFI$ | $\Delta TLI$ | $\Delta RMSEA$ | $\Delta SRMR$ | Decision |
| M1: Configural Invariance | 927.41 (202) | .986 | .983 | .021 (.019-.022) | .030 | — | — | — | — | — | — |
| M2: Metric Invariance | 975.47 (215)** | .985 | .983 | .021 (.019-.022) | .031 | 48.07 (13)** | -.001 | .000 | .000 | .001 | Accept |
| M3: Scalar Invariance | 1060.68 (228)** | .984 | .983 | .021 (.020-.022) | .031 | 85.21 (13)** | -.001 | .000 | .000 | .000 | Accept |
| **C. CAPE-9: Fathers v. adolescents** |  |  |  |  |  |  |  |  |  |  |  |
| Model | $\chi^2 (df)$ | CFI | TLI | RMSEA (95% CI) | SRMR | $\Delta\chi^2 (\Delta df)$ | $\Delta CFI$ | $\Delta TLI$ | $\Delta RMSEA$ | $\Delta SRMR$ | Decision |
| M1: Configural Invariance | 486.52 (48) | .980 | .970 | .020 (.018-.022) | .027 | — | — | — | — | — | — |
| M2: Metric Invariance | 580.49 (54)** | .976 | .968 | .020 (.019-.022) | .035 | 93.97 (6)** | -.004 | -.002 | .001 | .008 | Accept |
| M3: Scalar Invariance | 2263.07(60)** | .899 | .878 | .039 (.038-.041) | .059 | 1682.57 (6)** | -.077 | -.089 | .019 | .024 | Reject |
| M3a: Partial Scalar Invariance | 1037.97 (59)** | .955 | .945 | .026 (.025-.028) | .045 | 457.48 (5)** | -.021 | -.023 | .006 | .010 | Reject |

*Note.* Measurement invariance testing with nested model comparisons. CFI = comparative fit index; TLI = Tucker Lewis index (TLI); RMSEA = root mean square error of approximation; SRMR = standardized root mean square residual. Cut-off values used: .01 deviation cut-off for CFI and TLI, and .015 in RMSEA and .030 in SRMR for metric invariance; .015 in SRMR for scalar invariance. In the partial invariance model, we freed the intercept of item 4.
A. Boys $n = 8,717$ ; Girls $n = 10,118$
B. Pre-covid $n = 9,339$ ; During covid $n = 9,496$
C. Adolescents $n = 18,835$ ; Fathers $n = 28,793$
\* $p \leq .01$ , \*\* $p \leq .001$

**Table 2b.** Distress sub-scale: Test of measurement invariance across biological sex, pandemic status, and generations.

| **A. Boys v. girls** |  |  |  |  |  |  |  |  |  |  |  |
| --- | --- | --- | --- | --- | --- | --- | --- | --- | --- | --- | --- |
| Model | $\chi^2$ (df) | CFI | TLI | RMSEA (95% CI) | SRMR | $\Delta\chi^2$ (Δdf) | ΔCFI | ΔTLI | ΔRMSEA | ΔSRMR | Decision |
| M1: Configural Invariance | 785.95 (202) | .985 | .982 | .018 (.016-.019) | .034 | — | — | — | — | — | — |
| M2: Metric Invariance | 914.97 (215)** | .982 | .980 | .019 (.017-.020) | .037 | 129.02 (13)** | -.003 | -.002 | .001 | .003 | Accept |
| M3: Scalar Invariance | 1295.95(228)** | .973 | .971 | .022 (.021-.023) | .040 | 380.97 (13)** | -.009 | -.009 | .004 | .003 | Accept |
| **B. Pre- v. during COVID-19** |  |  |  |  |  |  |  |  |  |  |  |
| Model | $\chi^2$ (df) | CFI | TLI | RMSEA (95% CI) | SRMR | $\Delta\chi^2$ (Δdf) | ΔCFI | ΔTLI | ΔRMSEA | ΔSRMR | Decision |
| M1: Configural Invariance | 805.73 (202) | .986 | .984 | .018 (.017-.019) | .035 | — | — | — | — | — | — |
| M2: Metric Invariance | 901.08 (215)** | .984 | .982 | .018 (.017-.020) | .037 | 95.35 (13)** | -.002 | -.002 | .001 | .002 | Accept |
| M3: Scalar Invariance | 1003.24 (228)** | .982 | .981 | .019 (.018-.020) | .037 | 28.67 (13)** | -.002 | -.001 | .000 | .000 | Accept |
| **C. CAPE-9: Fathers v. adolescents** |  |  |  |  |  |  |  |  |  |  |  |
| Model | $\chi^2$ (df) | CFI | TLI | RMSEA (95% CI) | SRMR | $\Delta\chi^2$ (Δdf) | ΔCFI | ΔTLI | ΔRMSEA | ΔSRMR | Decision |
| M1: Configural Invariance | 306.49(48) | .985 | .977 | .015 (.013-.017) | .034 | — | — | — | — | — | — |
| M2: Metric Invariance | 338.65 (54)** | .983 | .978 | .015 (.013-.016) | .038 | 32.2 (6)* | -.002 | .000 | .000 | .004 | Accept |
| M3: Scalar Invariance | 1005.01 (60)** | .944 | .933 | .026 (.024-.027) | .051 | 666.37 (6)** | -.039 | -.045 | .011 | .013 | Reject |
| M3a: Partial Scalar Invariance | 568.36 (59) | .970 | .963 | .019 (.018-.020) | .042 | 229.71 (5)** | -.013 | -.015 | .004 | .004 | Reject |

*Note.* Measurement invariance testing with nested model comparisons. CFI = comparative fit index; TLI = Tucker Lewis index (TLI); RMSEA = root mean square error of approximation; SRMR = standardized root mean square residual. Cut-off values used: .01 deviation cut-off for CFI and TFI, and .015 in RMSEA and .030 in SRMR for metric invariance; .015 in SRMR for scalar invariance. In the partial invariance model, we freed the intercept of item 4.
A. Boys $n = 8,717$ ; Girls $n = 10,118$
B. Pre-covid $n = 9,339$ ; During covid $n = 9,496$
C. Adolescents $n = 18,835$ ; Fathers $n = 28,793$
\* $p \leq .01$ , \*\* $p \leq .001$

We found evidence for metric, but not scalar, invariance in CAPE-9 subscales across adolescents and fathers, indicating that while the factor structure and loadings were similar, the intercepts were not. Observing the modification indices suggested that freeing the equality constraint between groups for the *electrical influence* item intercept might improve model fit for both the frequency and distress scales. However, while freeing this parameter improved model fit, we were not able to establish partial scalar invariance (Tables 2a and 2b). Matched sample sizes suggested a negligible effect of unequal sample size on measurement invariance estimates (Supplementary Table S5).

### Associations between CAPE-16 sum scores and psychiatric diagnoses

Figure 2 shows associations between CAPE-16 sum scores and diagnoses in adolescents. Supplementary tables S6a and S6b provide detailed estimates.

**Figure 2.**
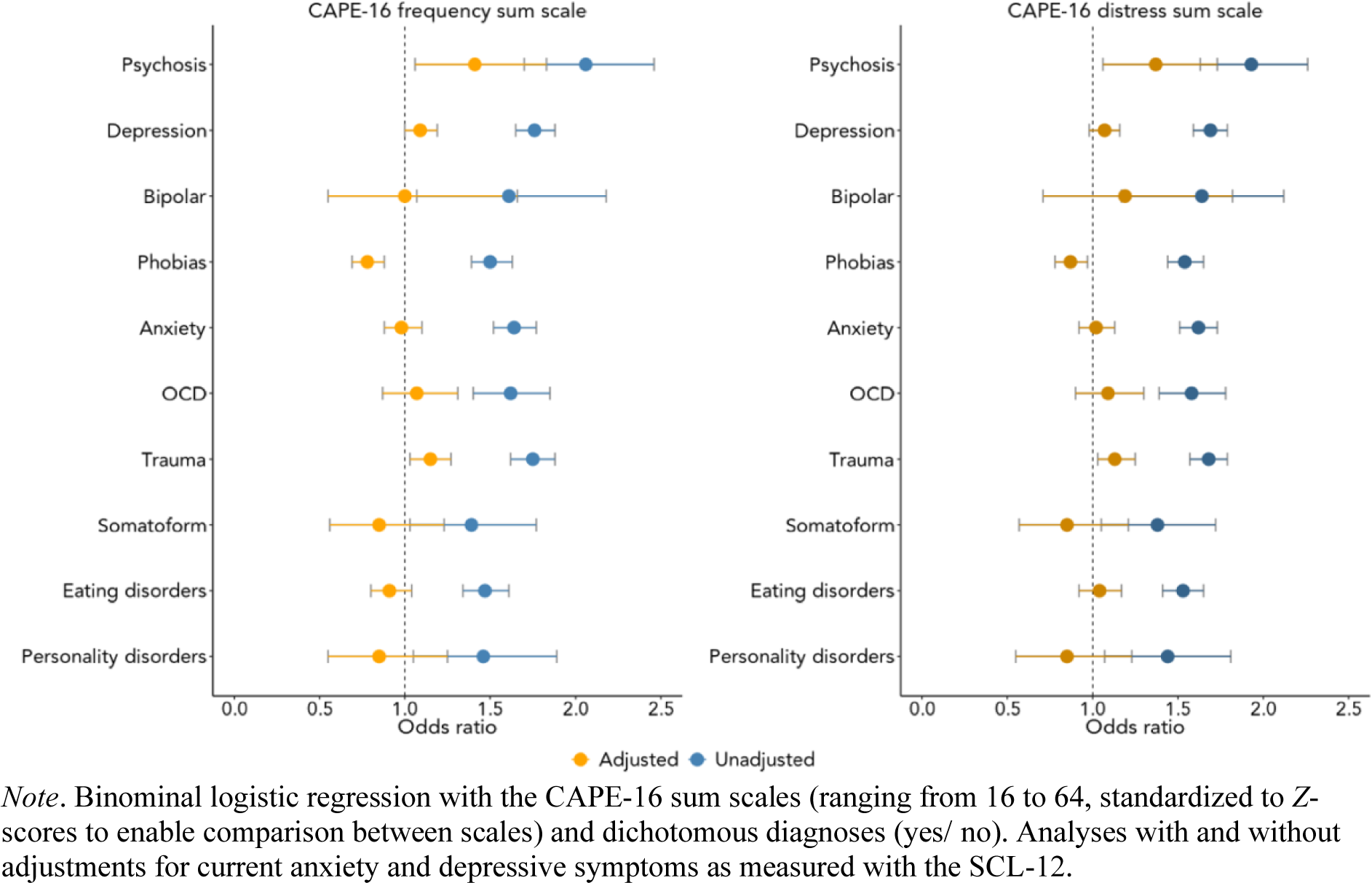
Associations between the CAPE-16 scales and psychiatric diagnoses – with and without adjustments for current anxiety and depressive symptoms

Both the frequency and distress scales were positively associated with all diagnoses but more with psychotic disorders (*frequency-psychosis:* OR = 2·06; 97·5% CI = 1·70–2·46; *distress-psychosis:* OR = 1·93; 97·5% CI = 1·63–2·26) (Figure 2). After adjusting for current emotional symptoms, the scales were only positively associated with psychotic disorders (*frequency*: OR = 1·41; 97·5% CI = 1·06–1·83; *distress*: OR = 1·37; 97·5% CI = 1·06–1·73) and negatively associated with phobias. Estimates from inverse probability weighted analyses were similar (Supplementary Tables S9a-b).

### Associations between CAPE-16 items and psychiatric diagnoses

Several frequency items were positively associated with one or more diagnoses (Table S7a). Notably, *hidden meaning* was positively associated with psychotic disorders (OR = 2·50; 97·5% CI = 1·42– 4·28), while e*lectrical influence* was negatively associated with several diagnoses.

Similarly, most distress items were positively associated with multiple diagnoses (Table S7b), with *beliefs about stalking* most associated with psychotic disorders (OR = 1·93; 97·5% CI = 1·18–3·07). *Electrical influence* was negatively associated with depression and anxiety. For both the frequency and distress scales, most item-specific associations disappeared after adjustment for current emotional symptoms, with some exceptions (Table S7a-b).

#### Exploratory analyses: Predicting psychosis using CAPE-16

Supplementary Figure 4 gives a summary of model performances, while Table S9 presents detailed results. Across the four models, average sensitivity ranged between 65·75-69·76 %, specificity between 82·39-85·37%, and balanced accuracy between 74·42-76·81%. Sensitivity, specificity, and balanced accuracies were statistically significant (at α = 0 · 05) in all models.

## Discussion

Our evaluation of the CAPE-16 questionnaire demonstrated good scale reliability and evidence for a previously observed three-factor structure^6^ in adolescents. The CAPE-16 scales were significantly associated with subsequent psychiatric diagnoses, particularly psychosis. There were small response differences across sex, before/during the COVID-19 pandemic, but larger differences across generations, specifically related to the *electrical influence* item. Together, these findings support CAPE-16 as a reliable tool for studying PEs in today’s youth.

Our measurement invariance testing indicated that scores were comparable across biological sex and pandemic status. This aligns with Barbosa et al. who found the CAPE positive scale to be equivalent across gender^28^. In our study, girls reported more PEs than boys, which is consistent with previous evidence that adolescent girls report more affective symptoms than boys^9^ but rather incongruent with the fact that boys are at a higher risk of psychosis^29^. One explanation may be that concurrent emotional symptoms have a greater impact on girls’ PE reports than boys’^19^. Further, those who responded during the COVID-19 pandemic reported slightly more frequent and distressing PEs than pre-pandemic respondents. This is in line with increased reported mental health issues after onset of the COVID-19 pandemic and underlines the assumed mental burden caused by the pandemic ^10^. Still, CAPE-16 was invariant across pandemic groups, suggesting that its psychometric properties were not significantly affected by this major psychosocial societal stressor. Consequently, the COVID-19 pandemic provides a unique opportunity to evaluate context-based differences in the properties and usefulness of a psychological measurement, which has not been possible since the Spanish flu pandemic.

MoBa adolescents reported substantially more PEs than MoBa fathers (33·4% and 2·0%). Previous studies have also indicated higher average prevalence of PEs in youths than adults^1–3^. Differences in self-reported PEs could be due to developmental processes in adolescents, impacting cognition and emotion^30^. Differences may also be due to generational aspects, such as increased openness around mental health in young people^31^. We observed metric but not scalar invariance of CAPE-9 across the two generations. This means that the adolescent generation respond differently to the father generation, not only in number of endorsed experiences. They may also interpret questions differently, complicating interpretation of the comparison. One item (*electrical influence*) showed large variability in estimated intercepts and reports (12·1% of adolescents vs. 0·4% of fathers) across generations. This item also demonstrated consistent negative associations with psychiatric diagnoses in adolescents but not fathers^7^. The phrasing of *electrical influence* (“Do you ever feel as if electrical devices can influence the way you think?”) could capture how digital natives, like today’s youth, perceive the impact modern, digital technology has on our daily lives. Hence, this item may measure generational differences not directly related to PEs, and thus not accurately capture pathological adolescent PEs. As such, we may need to revise such items to better reflect the day-to-day life of adolescents growing up in a digital world.

CAPE-16 sum scores were positively associated with all subsequent psychiatric diagnoses, particularly psychotic disorders. This is in line with previous observations^4^ and supports the validity of CAPE-16. Adjusting for current emotional symptoms made associations more specific to psychotic disorders in accordance with our previous study of MoBa fathers^7^. This suggests that CAPE-16 may capture emotional symptoms during reporting and that adjusting for these may improve predictive accuracy. No individual items were particularly associated with subsequent psychotic illness. As of 2022, all CAPE-16 respondents are younger than 23 years old. Given the typical onset of psychosis during early adulthood^32^, more cohort individuals are likely to develop psychotic disorders later, leading to an underestimation in the observed associations. Still, associations survived adjustments for emotional symptoms and, following our inverse probability assessments, did not appear to be substantially affected by attrition bias.

The aim of our prediction analysis was to help validate CAPE-16 as a measure of psychosis-related phenomena by demonstrating its ability to predict psychosis diagnoses beyond chance levels, even in a sample with low case prevalence. Here, predicting psychosis with reasonable accuracy provides further evidence supporting the validity of CAPE-16. The analysis was not expected to yield high predictive value but is a preliminary step towards using CAPE-16 in combination with other predictors to develop comprehensive and useful prediction models in the future.

The major strength of the current study is the large, well-defined, transgenerational sample with linkage to diagnostic registries, and questionnaires administered on both sides of the COVID-19 pandemic. Previously understudied, we investigated item-specific diagnostic associations, evaluating both frequency and distress of experiences. PE-related distress is often not considered although distressing PEs may hold clinical significance^33^.

The MoBa sample has several limitations, including loss to follow up of individuals with severe mental illness^34^. Still, inverse probability weighted estimates did not appear substantially affected by attrition bias. Second, online surveys typically yield higher reports than paper-pen surveys^35^. Similarly, using self-report instead of clinician-rated interviews may account for up to 20% of score variance^1^. Despite these variations, both measurement modes show similar associations^2^.

## Conclusions

In the current Norwegian cohort study, we found that the CAPE-16, a measure of self-reported psychotic experiences, showed strong psychometric properties through factor analyses, scale reliability tests, and measurement invariance across sex and response before or during the COVID-19 pandemic. We observed that the father and adolescent generations responded similarly but with certain differences to the questionnaire, warranting caution in interpreting responses across generations, particularly regarding questions involving digital technology. Additionally, adolescents who reported psychotic experiences at age 14 were more likely to later receive psychiatric diagnoses, specifically primary psychotic disorders. Our findings support the use of CAPE-16 in modern adolescents, even 20 years after the original questionnaire was created.

## Research in context

### Evidence before this study

We conducted a literature search in PubMed and Google Scholar for articles published between 2002 and 2024 on the Community Assessment of Psychic Experiences (CAPE; search terms: “CAPE” and “psychometric testing”), psychotic experiences in adolescents (“psychotic experiences” and “adolescents”), and associations between psychotic experiences and psychiatric diagnoses (“psychotic experiences” and “mental illness” or “psychiatric diagnoses”). The original CAPE, a widely used instrument to measure self-reported psychotic experiences in the general population, was developed over 20 years ago. A few studies suggested that children who reported psychotic experiences were more likely to develop adult-onset psychotic and non-psychotic mental illness. Still, prospective studies confirming relationships between adolescent psychotic experiences and subsequent psychiatric diagnoses are lacking. From previous research, adolescents appear to report more psychotic experiences than adults, and girls may report more experiences than boys. The last generations of adolescents grow up in a more digital world than to the parent generation, which may impact reporting. Such generational differences, as well as large societal changes, like the COVID-19 pandemic, may impact adolescents’ interpretation and response to instruments like the CAPE. No study has so far investigated this. By validating the CAPE-16 questionnaire in modern adolescents, we aimed to determine its effectiveness in capturing psychotic experiences across generations and contexts.

## Added value of this study

Our study directly addresses whether previously developed self-report tools still accurately access psychotic experienced in adolescents, the first generation growing up in a digital society. Our unique data on psychotic experiences in adolescents (*n* = 18,835) and fathers (*n* = 28,793) enabled transgenerational comparisons that show higher levels of psychotic experiences in the younger generation. Similarly, responses from adolescents both before and during the COVID-19 pandemic suggest that CAPE-16 is reliable across a major psychosocial stressor. To our knowledge, this is the first study to report psychotic experiences from generations before and after digital revolution, and before and during the COVID-19 pandemic.

## Implications of all the available evidence

Adolescents’ psychotic experiences are associated with subsequent psychiatric disorders and has potential to inform early mental health interventions in the future. The CAPE-16 exhibits robust psychometric properties across sex and pandemic status in modern adolescents. The current findings suggest differences in how adolescents and adults respond to the measure, and that the tool may be improved by adjusting items related to the digital world. This has potential implications for how researchers collect and compare self-reported experiences from individuals of different generations. More researchers should investigate whether currently used measures of mental health symptoms still hold up in the digital context adolescents now live in.

## Data Availability

Access to raw data can be requested via direct application to NIPH and MoBa.
For requirements and other information, see: https://www.fhi.no/en/ch/studies/moba/for-forskere-artikler/research-and-data-access/. The protocols and consent forms used for data collection can be
found here: https://www.fhi.no/en/ch/studies/moba/for-forskere-artikler/questionnaires-from-moba/.

https://www.fhi.no/en/ch/studies/moba/for-forskere-artikler/questionnaires-from-moba/

## Acknowledgments

The Norwegian Mother, Father and Child Cohort Study is supported by the Norwegian Ministry of Health and Care Services and the Ministry of Education and Research. We are grateful to all the participating families in Norway who take part in this on-going cohort study. We thank the Norwegian Institute of Public Health (NIPH) for generating high-quality genomic data. This research is part of the HARVEST collaboration, supported by the Research Council of Norway (#229624). We also thank the NORMENT Centre for providing genotype data, funded by the Research Council of Norway (#223273), Southeast Norway Health Authorities and Stiftelsen Kristian Gerhard Jebsen. We further thank the Center for Diabetes Research, the University of Bergen for providing genotype data and performing quality control and imputation of the data funded by the ERC AdG project SELECTionPREDISPOSED, Stiftelsen Kristian Gerhard Jebsen, Trond Mohn Foundation, the Research Council of Norway, the Novo Nordisk Foundation, the University of Bergen, and the Western Norway Health Authorities. The current work was performed on Services for sensitive data (TSD), University of Oslo, Norway, with resources from UNINETT Sigma2 - the National Infrastructure for High-Performance Computing and Data Storage in Norway. We want to thank Prof. Jim van Os for counseling during initial implementation of CAPE in MoBa.

## Data sharing

The code used for data management and analysis used in this study are available on request from the corresponding author. Access to raw data can be requested via direct application to NIPH and MoBa. For requirements and other information, see: https://www.fhi.no/en/ch/studies/moba/for-forskere-artikler/research-and-data-access/. The protocols and consent forms used for data collection can be found here: https://www.fhi.no/en/ch/studies/moba/for-forskere-artikler/questionnaires-from-moba/.

## Contributor statements

VB: As the primary author of this paper, I take responsibility for the content, methodology, and findings presented in the manuscript. I have actively contributed to the conceptualization and design of the study, analysis, interpretation of the results, and writing the manuscript. I declare that this work is original and has not been published elsewhere, and that all co-authors have been appropriately credited for their contributions.

PP and LH: We have contributed to the design of the study, data analyses and interpretation, as well as revision of the manuscript. We have access to all data in the study. We have reviewed and approved the final version of the manuscript for submission. Our contributions are accurately represented in the manuscript.

NRB, EF, PJ, OBS, ES, KMR, and MT: We have contributed to the conceptualization of the study, planning of analyses, and interpretation of results. We have access to all data in the study and have contributed with several waves of revisions and have approved the final version of the manuscript. Our contributions are accurately represented in the manuscript.

OAA, AH, and IES: As supervisors to the primary author and lead researchers, we have provided substantial input in the conceptualization and design of the study, securing funding, supervising the research team, and critically revising the manuscript. We have access to all data in the study and take responsibility for the integrity of the work as a whole and ensure that all co-authors’ contributions are appropriately acknowledged in the manuscript. We have approved the final version of the manuscript for submission and will be accountable for the accuracy and integrity of the work.

