## Supplementary material for "Youth Psychotic Experiences: Diagnostic Associations and Evaluation of the CAPE-16": 240408_Supplementary.docx

**Supplementary materials**

1. **Figure S1.** *Description of sample selection*
2. **Table S1a.** *Sociodemographic characteristics of adolescents who responded to CAPE-16*
3. **Table S1b.** *Sociodemographic comparison between adolescent* *non-respondents and respondents*
4. **Table S2a.** *Adolescents – Diagnostic counts: non-respondents and respondents*
5. **Table S2b.** *Differences in diagnostic counts between the mothers and fathers of adolescent respondents and non-respondents*
6. **Table S3** *CAPE-16 questions – self-reported lifetime psychotic experiences*
7. **Table S4.** *Distribution of psychotic experiences – responses to the frequency and distress CAPE-16 subscales in adolescents*
8. **Table S5.** *Matched sample sizes: test of measurement invariance across fathers and adolescents*
9. **Table S6a.** *CAPE-16, frequency subscale: associations between psychiatric diagnoses and the CAPE-16 frequency subscale*
10. **Table S6b.** *CAPE-16, distress subscale: associations between psychiatric diagnoses and the CAPE-16 distress subscale*
11. **Table S7a.** *CAPE-15 frequency items: Association between diagnoses and frequency items in adolescents*
12. **Table S7b.** *CAPE-16 distress items: Association between diagnoses and distress items in adolescents*
13. **Note 1: Inverse probability weighting**
    a. Table 8a. IPW: Associations between psychiatric diagnoses and the CAPE-16 frequency subscale with inverse probability weights
    b. Table 8b. IPW: Associations between psychiatric diagnoses and the CAPE-16 distress subscale with inverse probability weights
14. **Note 2. Exploring the predictive value of CAPE-16 on psychosis**
    a. **Figure S2.** *Machine learning schematic employed in this study*
    b. **Figure S3.** *Summary of machine learning model performances*
    c. **Table S9.** *Detailed results from the four CAPE-16 prediction models*
15. **References**

**Figure S1.** *Description of sample selection*

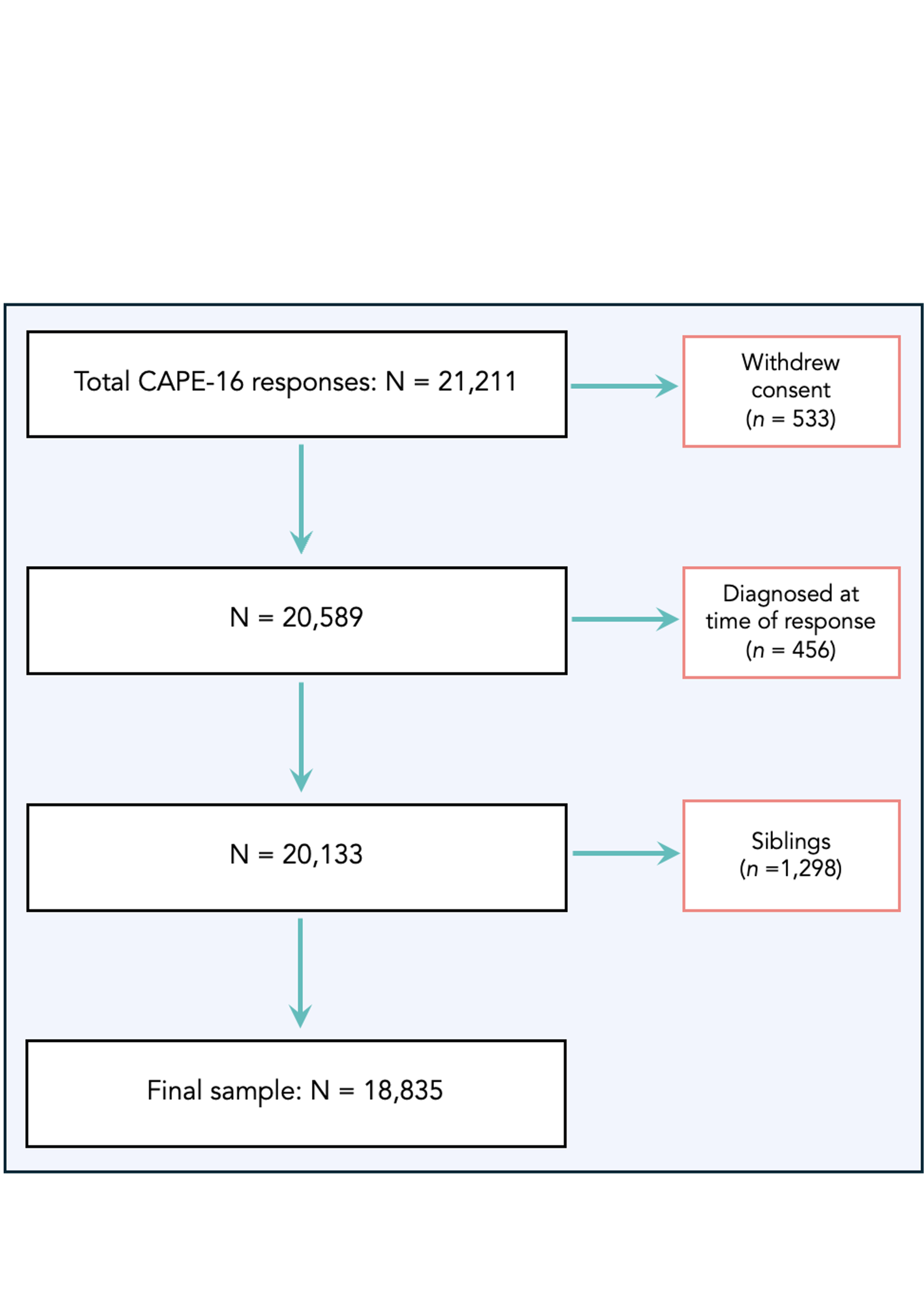

*Note.* Out of a total of 21,122 respondents, 533 individuals withdrew their consent at age 18. We further removed any individuals who were diagnosed with any of the included ICD-10 F-chapter diagnoses at time of response to CAPE-16 (*n* = 456). We then randomly removed one of each sibling pair to avoid relatedness among participants, resulting in a final sample of 18,835 individuals.

**Table S1a.** *Sociodemographic characteristics of adolescents who responded to CAPE-16*

| **Characteristic** | **Total**,  *n* = 18,835 | **Boys**,  *n* = 8,717 | **Girls**,  *n* = 10,118 | **Difference**^1^ | **95% CI**^12^ | ***p*-value**^1^ |
| --- | --- | --- | --- | --- | --- | --- |
| Age, *n* (%) |  |  |  | 0.01 | -0.02, 0.04 |  |
| 14 years | 11,245 (60%) | 5,180 (59%) | 6,065 (60%) |  |  |  |
| 15 years | 7,408 (39%) | 3,452 (40%) | 3,956 (39%) |  |  |  |
| 16 years | 200 (0.9%) | 94 (1.0%) | 106 (0.9%) |  |  |  |
| Lifetime use |  |  |  |  |  |  |
| Nicotine, *n* (%) | 2,674 (14%) | 1,453 (17%) | 1,221 (12%) | 4.6% |  | <0.001 |
| Alcohol, *n* (%) | 3,256 (17%) | 1,434 (17%) | 1,822 (18%) | -1.6% |  | 0.019 |
| Cannabis, *n* (%) | 264 (1.4%) | 155 (1.8%) | 109 (1.1%) | 0.70% |  | <0.001 |
| Drugs, other, *n* (%) | 238 (1.3%) | 126 (1.4%) | 112 (1.1%) | 0.34% |  | 0.12 |
| SCL-10, M (SD) | 15.2 (5.9) | 13.1 (4.1) | 16.9 (6.5) | -3.8 | -4.0, -3.7 | <0.001 |

^1^Standardized Mean Difference; 3-sample test for equality of proportions without continuity correction; Welch two sample t-test.

^2^CI = Confidence Interval; M = mean; SD = standard deviation.

**Table S1b.** *Sociodemographic comparisons between CAPE-16 non-respondents and respondents*

|  | **Non-respondents**,  *n* = 82,784 | **Respondent**,  *n* = 18,567 | ***p*-value**^2^ |
| --- | --- | --- | --- |
| Sex assigned at birth, *n* (%) |  |  | <0.001 |
| Not specified | 165 (0.2%) | 0 (0%) |  |
| Boy | 43,299 (52%) | 8,603 (46%) |  |
| Girl | 39,283 (47%) | 9,964 (54%) |  |
| Uncertain | 37 (<0.1%) | 0 (0%) |  |
| Parity, *n* (%) |  |  | 0.002 |
| First | 37,126 (45%) | 8,342 (45%) |  |
| Second | 29,659 (36%) | 6,440 (35%) |  |
| Third | 12,436 (15%) | 2,964 (16%) |  |
| Fourth | 2,709 (3.3%) | 643 (3.5%) |  |
| Fifth | 854 (1.0%) | 178 (1.0%) |  |
| Birthweight, gm, M (SD) | 3,546 (628) | 3,588 (570) | <0.001 |
| Missing | 238 | 7 |  |
| Mother’s age at birth, M (SD) | 30 (20) | 31 (19) | <0.001 |
| Father’s age at birth, M (SD) | 33 (23) | 34 (27) | <0.001 |
| Missing | 406 | 36 |  |
| Mother’s highest education*, *n* (%) |  |  | <0.001 |
| 9-year secondary school | 2,449 (3.1%) | 242 (1.4%) |  |
| 1-2 year high school | 4,335 (5.5%) | 567 (3.2%) |  |
| Vocational high school | 10,825 (14%) | 1,710 (9.7%) |  |
| 3-year high school | 12,087 (15%) | 2,177 (12%) |  |
| 4-year university/ college degree | 31,110 (40%) | 8,205 (46%) |  |
| University/ college, > 4 years | 17,718 (23%) | 4,779 (27%) |  |
| Missing | 4,260 | 887 |  |
| Father’s highest education*, *n* (%) |  |  | <0.001 |
| 9-year secondary school | 3,973 (5.3%) | 621 (3.7%) |  |
| 1-2 year high school | 4,744 (6.3%) | 783 (4.6%) |  |
| Vocational high school | 20,034 (27%) | 3,864 (23%) |  |
| 3-year high school | 9,503 (13%) | 1,929 (11%) |  |
| 4-year university/ college degree | 19,823 (26%) | 5,177 (31%) |  |
| University/ college, > 4 years | 17,169 (23%) | 4,562 (27%) |  |
| Missing | 7,538 | 1,631 |  |
| Mother’s income*, *n* (%) |  |  | <0.001 |
| No income | 2,129 (2.7%) | 339 (1.9%) |  |
| Under 150,000 NOK | 12,852 (16%) | 2,517 (14%) |  |
| 151,000-199,999 | 9,354 (12%) | 1,773 (9.8%) |  |
| 200,000-299,999 | 26,715 (34%) | 6,539 (36%) |  |
| 300,000-399,999 | 19,226 (24%) | 4,850 (27%) |  |
| 400,000-499,999 | 5,755 (7.2%) | 1,239 (6.9%) |  |
| Over 500,000 | 3,679 (4.6%) | 772 (4.3%) |  |
| Missing | 3,074 | 538 |  |
| Father’s income*, *n* (%) |  |  | <0.001 |
| No income | 820 (1.1%) | 139 (0.8%) |  |
| Under 150,000 NOK | 4,399 (5.7%) | 951 (5.4%) |  |
| 151,000-199,999 | 3,423 (4.4%) | 686 (3.9%) |  |
| 200,000-299,999 | 17,667 (23%) | 3,844 (22%) |  |
| 300,000-399,999 | 24,728 (32%) | 6,017 (34%) |  |
| 400,000-499,999 | 12,923 (17%) | 3,010 (17%) |  |
| Over 500,000 | 12,737 (16%) | 2,855 (16%) |  |
| Don’t know | 1,107 (1.4%) | 145 (0.8%) |  |
| Missing | 4,980 | 920 |  |
| Birth year, *n* (%) |  |  | <0.001 |
| 1999 | 46 (<0.1%) | 0 (0%) |  |
| 2000 | 1,997 (2.4%) | 0 (0%) |  |
| 2001 | 3,936 (4.8%) | 0 (0%) |  |
| 2002 | 7,878 (9.5%) | 410 (2.2%) |  |
| 2003 | 9,008 (11%) | 3,112 (17%) |  |
| 2004 | 9,258 (11%) | 3,829 (21%) |  |
| 2005 | 9,208 (11%) | 4,066 (22%) |  |
| 2006 | 11,980 (14%) | 4,906 (26%) |  |
| 2007 | 13,302 (16%) | 2,244 (12%) |  |

^1^Pearson's Chi-squared test; Wilcoxon rank sum test

^2^False discovery rate Benjamini-Hochberg corrected p-value

*During pregnancy 15 weeks of pregnancy

M = mean; SD = standard deviation

**Table S2a.** *Adolescents – Diagnostic counts: non-respondents and respondents*

| **Characteristic** | **Non-respondents**,  *n* = 95,072 | **Respondents**,  *n* = 18,835 | **p-value**^2^ |
| --- | --- | --- | --- |
| Any psychosis | 231 (0.2%) | 30 (0.2%) | 0.028 |
| Depressive episode | 3,133 (3.3%) | 510 (2.7%) | <0.001 |
| Bipolar | 170 (0.2%) | 18 (<0.1%) | 0.010 |
| Any developmental disability | 2,486 (2.6%) | 362 (1.9%) | <0.001 |
| Phobia | 2,675 (2.8%) | 339 (1.8%) | <0.001 |
| Anxiety | 1,031 (1.1%) | 90 (0.5%) | <0.001 |
| OCD | 3,684 (3.9%) | 351 (1.9%) | <0.001 |
| Stress- and trauma related disorders | 336 (0.4%) | 37 (0.2%) | <0.001 |
| Somatoform | 1,400 (1.5%) | 268 (1.4%) | 0.6 |
| Eating disorder | 280 (0.3%) | 31 (0.2%) | 0.002 |
| Personality disorder | 231 (0.2%) | 30 (0.2%) | 0.028 |

^1^Pearson's Chi-squared test; Wilcoxon rank sum test

^2^False discovery rate Benjamini-Hochberg corrected p-value

|  | **Mothers** | |  | **Fathers** | |  |
| --- | --- | --- | --- | --- | --- | --- |
| **Diagnoses** | **Non-respondent**,  N = 33,613 | **Respondent**,  N = 5,729 | **p-value**^1^ | **Non-respondent**,  N = 18,820 | **Respondent**,  N = 3,946 | **p-value**^1^ |
| Any organic mental disorders | 238 (0.7%) | 40 (0.7%) | >0.9 | 207 (1.1%) | 38 (1.0%) | 0.5 |
| Substance abuse, narcotics | 825 (2.5%) | 58 (1.0%) | <0.001 | 877 (4.7%) | 112 (2.8%) | <0.001 |
| Alcohol abuse | 968 (2.9%) | 81 (1.4%) | <0.001 | 1,284 (6.8%) | 204 (5.2%) | <0.001 |
| Any psychosis | 322 (1.0%) | 18 (0.3%) | <0.001 | 197 (1.0%) | 25 (0.6%) | 0.029 |
| Depressive episode | 7,282 (22%) | 1,127 (20%) | 0.001 | 3,140 (17%) | 582 (15%) | 0.006 |
| Bipolar | 1,224 (3.6%) | 149 (2.6%) | <0.001 | 579 (3.1%) | 102 (2.6%) | 0.14 |
| Any developmental disability | 33 (<0.1%) | 4 (<0.1%) | 0.6 | 24 (0.1%) | 1 (<0.1%) | 0.14 |
| Phobia | 3,875 (12%) | 527 (9.2%) | <0.001 | 879 (4.7%) | 134 (3.4%) | 0.001 |
| Anxiety | 5,371 (16%) | 800 (14%) | <0.001 | 1,901 (10%) | 339 (8.6%) | 0.008 |
| OCD | 579 (1.7%) | 78 (1.4%) | 0.065 | 210 (1.1%) | 41 (1.0%) | 0.7 |
| Stress- and trauma related disorders | 9,764 (29%) | 1,476 (26%) | <0.001 | 3,184 (17%) | 527 (13%) | <0.001 |
| Somatoform | 1,269 (3.8%) | 213 (3.7%) | 0.9 | 411 (2.2%) | 105 (2.7%) | 0.11 |
| Eating disorder | 943 (2.8%) | 119 (2.1%) | 0.002 | 45 (0.2%) | 8 (0.2%) | 0.7 |
| Personality disorder | 1,463 (4.4%) | 165 (2.9%) | <0.001 | 582 (3.1%) | 84 (2.1%) | 0.003 |
| Autism spectrum disorder | 83 (0.2%) | 16 (0.3%) | 0.7 | 72 (0.4%) | 12 (0.3%) | 0.5 |
| ADHD | 1,964 (5.8%) | 196 (3.4%) | <0.001 | 1,168 (6.2%) | 140 (3.5%) | <0.001 |

*Notes.* The table includes only those individuals who received psychiatric diagnosis from the Norwegian specialist health-care services between 2008-2022.

**Table S2b.** *Differences in diagnostic counts between the mothers and fathers of adolescent respondents and non-respondents*

^1^False discovery rate Benjamini-Hochberg corrected p-value

**Table S3** *CAPE-16 (CAPE -9 items in bold) – self-reported lifetime psychotic experiences*

| Factors | Abbreviations | The thoughts and feelings described here may seem unique to you, but they are more common than you might think*.  *Frequency.* How often have you been having these feelings or thoughts?  *Distress.* If you have experienced this, how affected are you by the experience? |
| --- | --- | --- |
| Persecutory ideation | **1. Delusions of reference** | Have you ever felt that what is printed in magazines and newspapers or said on TV specifically applies to you? |
|  | **2. Beliefs about stalking** | Do you ever feel as if you are being persecuted in any way? |
|  | **3. Beliefs about conspiracy** | Do you ever feel as if there is a conspiracy against you? |
|  | 10. Hidden meaning | Have you ever had the feeling as if people drop hints about you, or say things with a double meaning? |
|  | 11. Alienation | Do you ever feel as if some people are not what they seem to be? |
|  | 12. Odd looks | Do you ever feel that people look at you oddly because of your appearance? |
| Bizarre experiences | **4. Electrical influence** | Do you ever feel as if electrical devices can influence the way you think? |
|  | **5. Thought insertion** | Do you ever feel as if the thoughts in your head are not your own? |
|  | **6. Thought broadcasting** | Have your thoughts ever been so vivid that you were worried other people would hear them? |
|  | **7. External control** | Do you ever feel as if you are under the control of some force or power other than yourself? |
|  | 13. Thought withdrawal | Have you ever felt as if the thoughts in your head are being taken away from you? |
|  | 14. Thought-echo | Do you ever feel as if your own thoughts were being echoed back to you? |
|  | 16. Belief about impostor | Have you ever felt as if a double has taken the place of a family member, a friend or an acquaintance? |
| Perceptual abnormalities | **8. Hearing voices** | Have you ever heard voices when you were completely alone (not radio or TV)? |
|  | **9. Visual hallucinations** | Do you ever see objects, people, or animals that other people cannot see? |
|  | 15. Hearing voices talk | Do you ever hear voices talking to each other when you are alone? |

*Note.* Numbers after the abbreviations indicate the sequence in which the questions were ordered in the original MoBa questionnaire. The items in bold make up the CAPE-9 scale. The three factors are based on previous factor analyses conducted on the CAPE-15 ^1,2^ which we replicate for CAPE-16 in MoBa adolescents using confirmatory factor analysis.

**Table S4.** *Distribution of psychotic experiences – responses to the frequency and distress CAPE-16 subscales in adolescents*

| Frequency of experiences | Never | Sometimes | Often | Nearly always |  | Associated distress | Not at all | A little | Quite a lot | Very much |
| --- | --- | --- | --- | --- | --- | --- | --- | --- | --- | --- |
| Delusions of reference | 62.9 % | 33.5 % | 3.1 % | 0.5 % |  | Delusions of reference | 86.6 % | 12 % | 1.2 % | 0.2 % |
| Beliefs about stalking | 78.1 % | 18.7 % | 2.6 % | 0.6 % |  | Beliefs about stalking | 84.7 % | 11.8 % | 2.7 % | 0.8 % |
| Beliefs about conspiracy | 66.3 % | 29.3 % | 3.7 % | 0.6 % |  | Beliefs about conspiracy | 75 % | 18.1 % | 5.4 % | 1.5 % |
| Electrical influence | 54.2 % | 33.7 % | 10.4 % | 1.7 % |  | Electrical influence | 72.9 % | 22.1 % | 4.2 % | 0.8 % |
| Thought insertion | 74.6 % | 20.5 % | 4.1 % | 0.8 % |  | Thought insertion | 81.5 % | 13.5 % | 3.8 % | 1.2 % |
| Thought broadcasting | 67.5 % | 23.2 % | 7.4 % | 1.8 % |  | Thought broadcasting | 80.5 % | 14.4 % | 4 % | 1.1 % |
| External control | 83.9 % | 12.7 % | 2.6 % | 0.8 % |  | External control | 89.5 % | 7.4 % | 2.4 % | 0.8 % |
| Hearing voices | 83.5 % | 14 % | 1.9 % | 0.5 % |  | Hearing voices | 88.9 % | 8 % | 2.2 % | 0.9 % |
| Visual hallucinations | 91.2 % | 7.7 % | 0.9 % | 0.3 % |  | Visual hallucinations | 94.3 % | 4 % | 1.2 % | 0.5 % |
| Hidden meaning | 72.5 % | 23.3 % | 3.5 % | 0.6 % |  | Hidden meaning | 84.9 % | 11.8 % | 2.6 % | 0.7 % |
| Alienation | 63.9 % | 28.6 % | 6.5 % | 1.1 % |  | Alienation | 78.5 % | 15.8 % | 4.5 % | 1.2 % |
| Odd looks | 58.2 % | 31.6 % | 7.7 % | 2.5 % |  | Odd looks | 69.4 % | 18.5 % | 8.5 % | 3.6 % |
| Thought withdrawal | 90.2 % | 8.3 % | 1.2 % | 0.3 % |  | Thought withdrawal | 93 % | 5 % | 1.5 % | 0.5 % |
| Thought echoing | 83.4 % | 12.7 % | 2.8 % | 1.1 % |  | Thought echoing | 93 % | 5.4 % | 1.2 % | 0.4 % |
| Hearing voices talk | 93.2 % | 5.5 % | 0.9 % | 0.4 % |  | Hearing voices talk | 96.3 % | 2.7 % | 0.7 % | 0.3 % |
| Beliefs about impostor | 95.6 % | 3.9 % | 0.4 % | 0.1 % |  | Beliefs about impostor | 97 % | 2 % | 0.7 % | 0.3 % |

**Table S5.** *Matched sample sizes: test of measurement invariance across fathers and adolescents*

| **Frequency scale** |  |  |  |  |  |  |  |  |  |  |  |
| --- | --- | --- | --- | --- | --- | --- | --- | --- | --- | --- | --- |
| Model | χ^2^ (*df*) | CFI | TLI | RMSEA (95% CI) | SRMR | Δχ^2^ (Δ*df*) | ΔCFI | ΔTLI | ΔRMSEA | ΔSRMR | Decision |
| M1: Configural Invariance | 900.25 (48) | .989 | . 983 | .027(.026-.029) | .043 | **–** | **–** | **–** | **–** |  | **–** |
| M2: Metric Invariance | 1076.07 (54)** | .986 | .982 | .028(.027-.030) | .047 | 175.8 (6)** | -.002 | -.001 | .001 | .005 | Accept |
| M3: Scalar Invariance | 1524.93 (60)** | .981 | .980 | .030(.028-.031) | .051 | 448.86 (6)** | -.006 | -.002 | .002 | .004 | Accept |
| **Distress scale** |  |  |  |  |  |  |  |  |  |  |  |
| Model | χ2 (df) | CFI | TLI | RMSEA (95% CI) | SRMR | Δχ2 (Δdf) | ΔCFI | ΔTLI | ΔRMSEA | ΔSRMR | Decision |
| M1: Configural Invariance | 296.73 (48) | .985 | . 977 | .017(.015-.018) | .035 | – | – | – | – |  | – |
| M2: Metric Invariance | 320.42 (54)** | .984 | .979 | .016(.014-.018) | .040 | 23.69 (6)* | -.001 | .001 | .000 | .006 | Accept |
| M3: Scalar Invariance | 950.54 (60)** | .946 | .935 | .028(.027-0.030) | .053 | 630.12 (6)** | -.038 | -.043 | .012 | .012 | Reject |

*Note.* Measurement invariance testing with nested model comparisons. CFI = comparative fit index; TLI = Tucker Lewis index (TLI); RMSEA = root mean square error of approximation; SRMR = standardized root mean square residual. Cut-off values used: -.01 deviation cut-off for CFI and TFI, and .015 in RMSEA and .030 in SRMR for metric invariance; .015 in SRMR for scalar invariance.

N = 37,670: Adolescents *n* = 18,835; Fathers *n* = 18,835.

* *p ≤* .01, ** *p ≤* .001

Table S6a. CAPE-16, frequency subscale: associations between psychiatric diagnoses and the CAPE-16 frequency subscale

|  | Not adjusted for SCL-10 | | | | Adjusted for SCL-10 | | | |
| --- | --- | --- | --- | --- | --- | --- | --- | --- |
| Diagnostic variables | OR | Lower CI  (2.5%) | Upper CI  (97.5%) | *p*-value^1^ | OR | Lower CI  (2.5%) | Upper CI  (97.5%) | *p*-value^1^ |
| Any psychosis | 2.06 | 1.70 | 2.46 | 1.04E-14 | 1.41 | 1.06 | 1.83 | 0.031 |
| Depressive episode | 1.76 | 1.65 | 1.88 | 9.78E-66 | 1.09 | 1.00 | 1.19 | 0.153 |
| Bipolar disorder | 1.61 | 1.07 | 2.18 | 0.009 | 1.00 | 0.55 | 1.66 | 0.986 |
| Phobias | 1.50 | 1.39 | 1.63 | 1.52E-23 | 0.78 | 0.69 | 0.88 | >0.001 |
| Anxiety disorders | 1.64 | 1.52 | 1.77 | 2.09E-35 | 0.98 | 0.88 | 1.10 | 0.861 |
| OCD | 1.62 | 1.40 | 1.85 | 7.67E-12 | 1.07 | 0.87 | 1.31 | 0.607 |
| Trauma- and stress related disorders | 1.75 | 1.62 | 1.88 | 7.33E-49 | 1.15 | 1.03 | 1.27 | 0.046 |
| Somatoform | 1.39 | 1.03 | 1.77 | 0.017 | 0.85 | 0.56 | 1.23 | 0.607 |
| Eating disorders | 1.47 | 1.34 | 1.61 | 3.21E-16 | 0.91 | 0.80 | 1.04 | 0.349 |
| Personality disorders | 1.46 | 1.05 | 1.89 | 0.011 | 0.85 | 0.55 | 1.25 | 0.607 |

*Note*. Binominal logistic regression with a continuous CAPE-16 frequency subscale (ranging from 16 to 64, standardized to z-scores to enable comparison between scales) and dichotomous diagnoses (yes/ no). Analyses with and without adjustments for current anxiety and depressive symptoms as measured with the SCL-12.

^1^False discovery rate Benjamini-Hochberg corrected p-value.

Table S6b: CAPE-16, distress subscale: associations between psychiatric diagnoses and the CAPE-16 distress subscale

|  | Not adjusted for SCL-10 | | | | Adjusted for SCL-10 | | | |
| --- | --- | --- | --- | --- | --- | --- | --- | --- |
| Diagnostic variables | OR | Lower CI  (2.5%) | Upper CI  (97.5%) | *p*-value^1^ | OR | Lower CI  (2.5%) | Upper CI  (97.5%) | *p*-value^1^ |
| Any psychosis | 1.93 | 1.63 | 2.26 | 8.33E-16 | 1.37 | 1.06 | 1.73 | 0.041 |
| Depressive episode | 1.69 | 1.59 | 1.79 | 2.38E-67 | 1.07 | 0.98 | 1.16 | 0.333 |
| Bipolar disorder | 1.64 | 1.17 | 2.12 | 0.001 | 1.19 | 0.71 | 1.82 | 0.555 |
| Phobias | 1.54 | 1.44 | 1.65 | 5.6E-34 | 0.87 | 0.78 | 0.97 | 0.041 |
| Anxiety disorders | 1.62 | 1.51 | 1.73 | 1.76E-41 | 1.02 | 0.92 | 1.13 | 0.684 |
| OCD | 1.58 | 1.39 | 1.78 | 2.6E-13 | 1.09 | 0.9 | 1.3 | 0.555 |
| Trauma- and stress related disorders | 1.68 | 1.57 | 1.79 | 1.21E-51 | 1.13 | 1.03 | 1.25 | 0.041 |
| Somatoform | 1.38 | 1.05 | 1.72 | 0.008 | 0.85 | 0.57 | 1.21 | 0.555 |
| Eating disorders | 1.53 | 1.41 | 1.65 | 1.08E-25 | 1.04 | 0.92 | 1.17 | 0.555 |
| Personality disorders | 1.44 | 1.07 | 1.81 | 0.007 | 0.85 | 0.55 | 1.23 | 0.555 |

*Note*. Binominal logistic regression with a continuous CAPE-16 distress subscale (ranging from 16 to 64, standardized to z-scores to enable comparison between scales) and dichotomous diagnoses (yes/ no). Analyses with and without adjustments for current anxiety and depressive symptoms as measured with the SCL-12.

^1^False discovery rate Benjamini-Hochberg corrected p-value.

**Table S7a.** CAPE-16 frequency items: Association between diagnoses and frequency items in adolescents

| Not adjusted for SCL-10 | | | | | | Adjusted for SCL-10 | | | |
| --- | --- | --- | --- | --- | --- | --- | --- | --- | --- |
| Diagnoses | Items | OR | Lower CI  (2.5%) | Upper CI  (97.5%) | *p*-value^1^ | OR | Lower CI  (2.5%) | Upper CI  (97.5%) | *p*-value^1^ |
| Psychosis | Hidden meaning | 2.50 | 1.42 | 4.28 | 0.014 |  |  |  |  |
| Depression | Beliefs about stalking | 1.32 | 1.13 | 1.54 | 0.007 |  |  |  |  |
|  | Electrical influence | 0.70 | 0.60 | 0.80 | >0.001 | 0.70 | 0.60 | 0.81 | >0.001 |
|  | Thought insertion | 1.41 | 1.20 | 1.65 | 0.001 |  |  |  |  |
|  | Odd looks | 1.84 | 1.62 | 2.08 | >0.001 | 1.33 | 1.16 | 1.52 | >0.001 |
| Bipolar disorder |  |  |  |  |  |  |  |  |  |
| Phobias | Electrical influence | 0.75 | 0.63 | 0.88 | 0.01 | 0.75 | 0.64 | 0.89 | 0.032 |
|  | Odd looks | 2.16 | 1.87 | 2.5 | >0.001 | 1.36 | 1.16 | 1.58 | >0.001 |
| Anxiety disorders | Electrical influence | 0.75 | 0.63 | 0.89 | 0.012 | 0.77 | 0.65 | 0.91 | 0.046 |
|  | Thought insertion | 1.41 | 1.16 | 1.71 | 0.009 |  |  |  |  |
|  | Odd looks | 1.53 | 1.3 | 1.78 | >0.001 |  |  |  |  |
| OCD | Thought insertion | 1.88 | 1.31 | 2.65 | 0.007 |  |  |  |  |
| Trauma | Odd looks | 1.62 | 1.39 | 1.88 | >0.001 |  |  |  |  |
| Eating disorders | Thought insertion | 1.38 | 1.11 | 1.71 | 0.034 |  |  |  |  |
|  | External control | 1.40 | 1.12 | 1.73 | 0.025 |  |  |  |  |
|  | Odd looks | 1.81 | 1.53 | 2.14 | >0.001 | 1.36 | 1.14 | 1.63 | 0.032 |
| Personality disorders |  |  |  |  |  |  |  |  |  |

***Note.*** Results from multiple logistic regression analyses showing the associations between CAPE-16 frequency items and psychiatric diagnoses, with and without adjustments for current anxiety and depressive symptoms (SCL-10). P-values were adjusted for multiple tests by controlling for the false discovery rate. To improve readability, non-significant results were removed (*p* >= .05)

**Table S7b.** CAPE-16 distress items: Association between diagnoses and distress items in adolescents

| Not adjusted for SCL-10 | | | | | | Adjusted for SCL-10 | | | |
| --- | --- | --- | --- | --- | --- | --- | --- | --- | --- |
| Diagnoses | Items | OR | Lower CI  (2.5%) | Upper CI  (97.5%) | *p*-value^1^ | OR | Lower CI  (2.5%) | Upper CI  (97.5%) | *p*-value^1^ |
| Psychosis | Beliefs about stalking | 1.93 | 1.18 | 3.07 | 0.049 |  |  |  |  |
| Depression | Beliefs about stalking | 1.29 | 1.11 | 1.48 | 0.012 |  |  |  |  |
|  | Electrical influence | 0.71 | 0.60 | 0.82 | >0.001 | 0.66 | 0.55 | 0.78 | >0.001 |
|  | Thought insertion | 1.30 | 1.13 | 1.49 | >0.001 |  |  |  |  |
|  | External control | 1.33 | 1.14 | 1.54 | >0.001 |  |  |  |  |
|  | Odd looks | 1.40 | 1.26 | 1.56 | >0.001 | 1.21 | 1.07 | 1.36 | 0.08 |
| Bipolar disorder | Thought broadcasting | 2.87 | 1.59 | 4.97 | >0.001 |  |  |  |  |
| Phobias | Beliefs about stalking | 1.27 | 1.07 | 1.50 | 0.046 |  |  |  |  |
|  | Thought broadcasting | 1.25 | 1.06 | 1.46 | 0.046 |  |  |  |  |
|  | Odd looks | 1.58 | 1.39 | 1.79 | >0.001 | 1.24 | 1.08 | 1.43 | 0.07 |
| Anxiety disorders | Electrical influence | 0.74 | 0.61 | 0.89 | 0.021 | 0.72 | 0.58 | 0.87 | 0.08 |
|  | Thought insertion | 1.55 | 1.31 | 1.83 | >0.001 |  |  |  |  |
| OCD | Thought insertion | 1.75 | 1.28 | 2.37 | >0.001 |  |  |  |  |
| Trauma | Beliefs about stalking | 1.29 | 1.09 | 1.53 | 0.025 |  |  |  |  |
|  | Beliefs about conspiracy | 1.27 | 1.09 | 1.47 | 0.021 |  |  |  |  |
|  | Visual hallucinations | 1.35 | 1.10 | 1.64 | 0.025 |  |  |  |  |
|  | Odd looks | 1.22 | 1.07 | 1.39 | 0.025 |  |  |  |  |
| Somatoform disorder | Beliefs about stalking | 2.13 | 1.31 | 3.31 | 0.012 | 2.24 | 1.29 | 3.72 | 0.08 |
| Eating disorders | Delusions of reference | 1.41 | 1.12 | 1.77 | 0.025 | 1.42 | 1.10 | 1.80 | 0.09 |
|  | External control | 1.47 | 1.20 | 1.77 | >0.001 |  |  |  |  |
|  | Odd looks | 1.48 | 1.28 | 1.71 | >0.001 | 1.27 | 1.08 | 1.49 | 0.09 |
| Personality disorders |  |  |  |  |  |  |  |  |  |

***Note.*** Results from multiple logistic regression analyses showing the associations between CAPE-16 distress items and psychiatric diagnoses, with and without adjustments for current anxiety and depressive symptoms (SCL-10). P-values were adjusted for multiple tests by controlling for the false discovery rate. To improve readability, non-significant results were removed (*p* >= .05).

**Note 1. Inverse probability weighting**

Continued participation in cohort studies, like MoBa^3,4^, is associated with certain sociodemographic and psychiatric characteristics in those who continue to respond^4^. To test the effect of attrition bias on our associations, we investigated the impact of including inverse probability weights (IPW^5^) based on differences between CAPE non-/respondents in our regression analyses. In IPW, the first step is to predict continued participation based on certain variables related to participation by creating scores that indicated the probability that each participant responded to the follow-up or not. Secondly, these probabilities are weighted by the inverse probability of response. The resulting weights are added to the exposure-outcome analyses of interest, thereby inflating the effect of under-represented observations^4–6^.

Metten et al.^6^ argue that weights will not improve attrition bias if the variables chosen are not also related to the associations that are being investigated. Because of this, we specifically chose to test the predictive effect of variables that are both believed to be related to the continued participation in MoBa and associations between CAPE-16 and psychiatric diagnoses. We generated propensity scores (estimates of the probability that a person responded to the 14-year questionnaires based on the variables we selected) by running logistic regression models. We then compared multiple propensity score models on their ability to predict response through area under the curve (AUC) analyses. The model that best predicted whether a person would respond to the 14-year questionnaires included: Biological sex, mother’s and father’s age at birth, mother’s education, psychiatric diagnoses (of both parents and children; please see Table S2b and S2c) in which the prevalence differed significantly between CAPE-16 respondents and non-respondents (substance use, anxiety disorders , phobias, trauma-related disorders, ASD, and ADHD). The AUC score of this model was 0.604.

Based on the propensity scores, we generated IPW-values, and re-ran our CAPE-diagnoses analyses (Supplementary Tables 9a to d) and compared the results with the unweighted analyses.

Table S8a. Inverse probability weighting - CAPE-16, frequency subscale: associations between psychiatric diagnoses and the CAPE-16 frequency subscale with inverse probability weights

| Diagnostic variables | OR | Lower CI  (2.5%) | Upper CI  (97.5%) | *p*-value^1^ |
| --- | --- | --- | --- | --- |
| Any psychosis | 2.02 | 1.65 | 2.43 | 1.29E-12 |
| Depressive episode | 1.72 | 1.61 | 1.84 | 6.15E-54 |
| Bipolar disorder | 1.49 | 0.97 | 2.08 | 0.042 |
| Phobias | 1.46 | 1.34 | 1.58 | 5.25E-18 |
| Anxiety disorders | 1.61 | 1.49 | 1.75 | 1.6E-30 |
| OCD | 1.57 | 1.35 | 1.8 | 1.55E-09 |
| Trauma- and stress related disorders | 1.74 | 1.59 | 1.9 | 1.8E-33 |
| Somatoform | 1.39 | 1.03 | 1.78 | 0.021 |
| Eating disorders | 1.47 | 1.33 | 1.62 | 7.93E-15 |
| Personality disorders | 1.51 | 1.09 | 1.95 | 0.006 |

*Note.* Results from multiple logistic regression analyses estimating the associations between the CAPE-16 frequency subscale and psychiatric diagnoses with inverse probability weights (IPW) added.

^1^False discovery rate Benjamini-Hochberg corrected p-value.

Table S8b. IPW - CAPE-16, distress subscale: associations between psychiatric diagnoses and the CAPE-16 frequency subscale with inverse probability weights

| Diagnostic variables | OR | Lower CI  (2.5%) | Upper CI  (97.5%) | *p*-value^1^ |
| --- | --- | --- | --- | --- |
| Any psychosis | 1.94 | 1.63 | 2.28 | 1.66E-14 |
| Depressive episode | 1.66 | 1.56 | 1.77 | 4.71E-57 |
| Bipolar disorder | 1.58 | 1.10 | 2.08 | 0.004 |
| Phobias | 1.51 | 1.4 | 1.62 | 1.06E-27 |
| Anxiety disorders | 1.61 | 1.49 | 1.72 | 2.24E-37 |
| OCD | 1.55 | 1.36 | 1.75 | 2.91E-11 |
| Trauma- and stress related disorders | 1.72 | 1.58 | 1.86 | 5.21E-39 |
| Somatoform | 1.38 | 1.05 | 1.73 | 0.011 |
| Eating disorders | 1.53 | 1.41 | 1.66 | 1.34E-23 |
| Personality disorders | 1.47 | 1.09 | 1.85 | 0.004 |

*Note.* Results from multiple logistic regression analyses estimating the associations between the CAPE-16 distress subscale and psychiatric diagnoses with inverse probability weights (IPW) added.

^1^False discovery rate Benjamini-Hochberg corrected p-value.

**Note 2: Exploring the predictive value of CAPE-16 on psychosis**

To create the model where we predict psychosis using CAPE-16, we specified a reproducible *template tree* and then passed this tree to the *fitcensemble* function in MATLAB with the following parameters – ‘method’ as ‘RUSBoost’^7^, ‘LearnRate’ as ‘0.1’, and ‘NumLearningCycles’ as ‘100’. Analyses were conducted in MATLAB R2023a (The MathWorks, Natick, USA).

To get a reliable estimate of model performance, we performed 50 repeats of 5-fold stratified cross-validation (i.e., each fold would have 24 or 25 adolescents with psychosis for model training and 6 or 7 adolescents with psychosis for testing; this process was repeated 50 times). For every repeat, we calculated the average (over the five folds) sensitivity, specificity, and balanced accuracy (defined as the average of sensitivity and specificity); then, we averaged these values over the 50 repeats to get the mean and standard deviations of the classifier *test* performance. It is important to highlight the use of balanced accuracy in case of skewed distribution of categorical outcome variables. Consider, for example, a trivial classifier that always predicts “not psychosis” as the output – in this case, the accuracy would be ~99.8% (with 0% sensitivity and 100% specificity; the balanced accuracy in this case will be 50%). Finally, to evaluate whether our classifier performance was above chance, we employed permutation testing as described in Ojala and Garriga^8.^ Briefly, we shuffled the labels (psychosis or not psychosis) and then retrained the classifier using the same settings as in the non-permuted case. We performed 100 permutations of 5-fold cross-validation and then computed the *p*-value as the fraction of times the permutation performance (sensitivity, specificity, and balanced accuracy) became equal to or exceeded the actual average (over 50 repeats) performance. Note that the usual practice in this case is to add a value of 1 in both the numerator and denominator; therefore, the smallest *p*-value in our case would be $\frac{1}{101}=0.0099$.

Since we were interested in examining the predictive value of CAPE-16 questionnaire response items, which might be confounded by individuals’ current emotional and behavioral symptoms, we created another model where we regressed out the effect of SCL-10 from each of the 48 CAPE questionnaires features – this regression was carried out within cross-validation (i.e., for each fold, we *learned* the regression parameters using the training set and then *applied* the regression parameters to the test set, thereby avoiding information leakage). Overall, we ran four models: two with the complete sample (with and without regression of SCL-10 variables), and two where any ICD-10 F chapter diagnoses were removed from subjects who did not have psychosis (with and without regression of SCL-10 variables). The schematic for the overall machine learning pipeline is shown in Supplementary Figure 3, detailed estimates are shown in Supplementary Figure 4 and Supplementary Table 10.

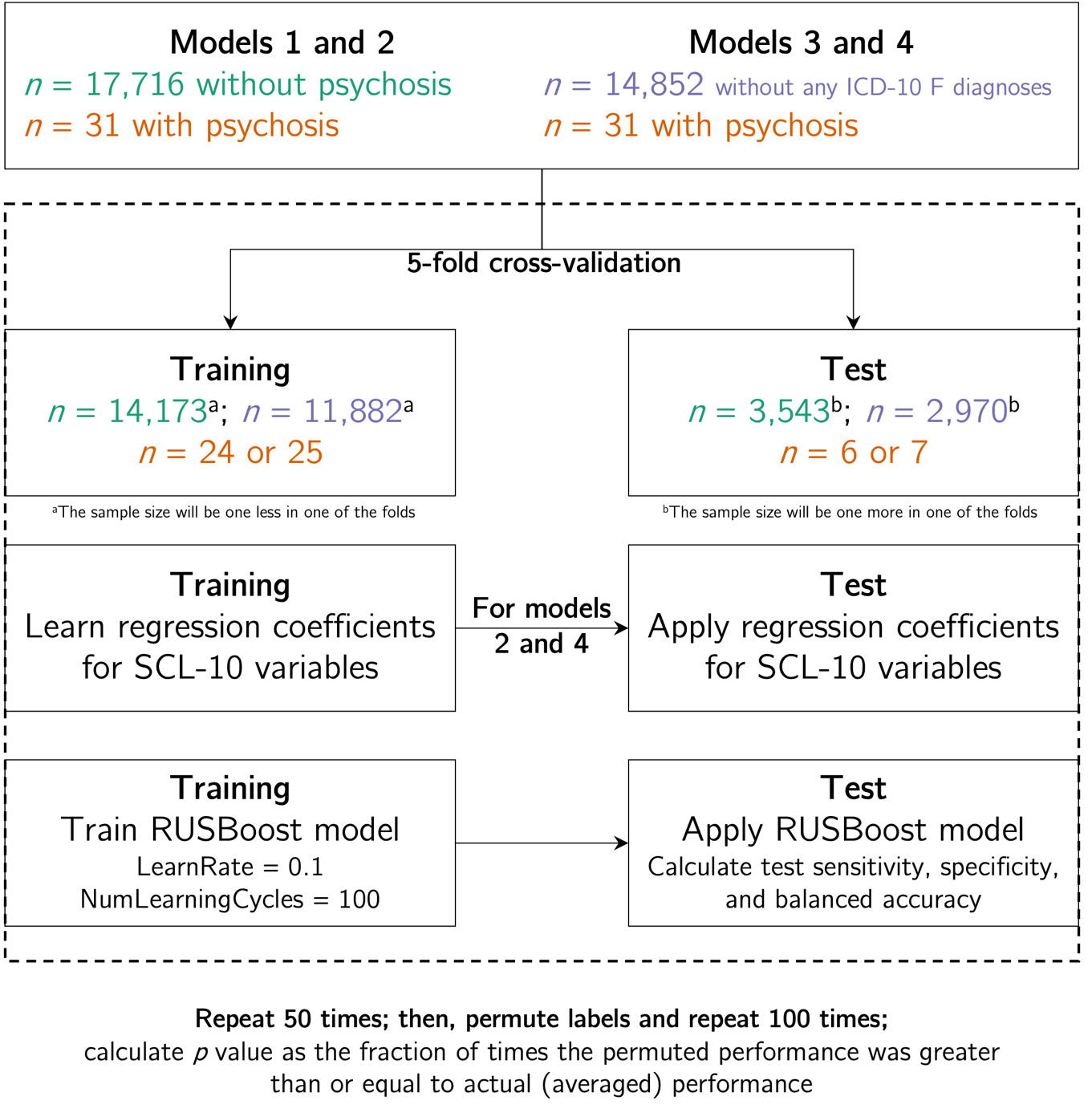

**Figure 2:** Machine learning schematic employed in this study. We trained and tested four models; in each of the model there were 31 adolescents that had a diagnosis of psychosis. For models 1 and 2, there were 17,716 individuals without psychosis while for models 3 and 4, we eliminated data of any adolescent (without psychosis) who had any ICD-10 F chapter diagnoses, resulting in 14,852 individuals. We performed 50 repeats of 5-fold cross validation where we trained and tested a RUSBoost model. Additionally, for models 2 and 4, we regressed the effect of SCL-10 variables from the 48 CAPE-16 features (within cross-validation framework) prior to training and testing the RUSBoost model. In all scenarios, we calculated the test sensitivity, specificity, and balanced accuracy (average of sensitivity and specificity). To evaluate whether our models performed above chance levels, we permuted the class labels and repeated the entire 5-fold cross-validation process 100 times. Then, we calculated the *p*-value as the fraction of times the permuted performance was greater than or equal to the actual average performance.

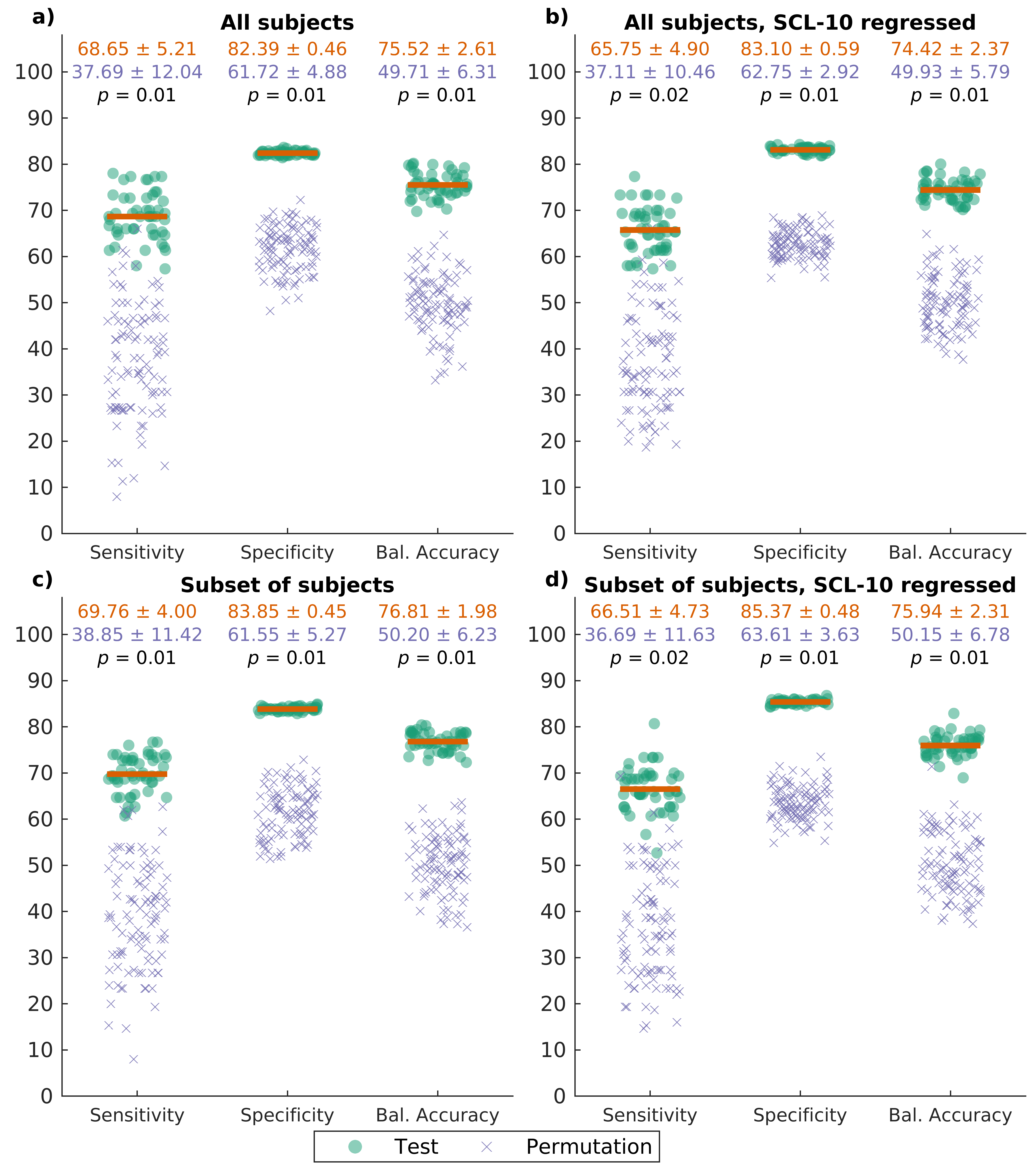

**Figure 3:** Summary of machine learning model performances. For each of the four panels, the *x*-axis shows sensitivity, specificity, and balanced accuracy (defined as the average between sensitivity and specificity) while the *y*-axis shows the value in percentages. The green dots indicate the (average over 5-fold cross-validation) test performance of the model (50 repeats) while the purple crosses indicate the (average over 5-fold cross-validation) permutation test performance of the model (100 repeats). The solid orange line indicates the average (over 50 repeats) for the test set. The numbers on the top of each panel indicate the mean and standard deviations for the test set (orange text), permutation set (purple text), and the *p*-value (black text) computed as the fraction of times the permutation performance became equal to or greater than the test performance. Panel **a)** shows the performance of the model that uses the full dataset (excluding subjects with missing values), panel **b)** shows the performance of the model that uses the full dataset (excluding subjects with missing values) where we additionally adjusted the CAPE-16 features for current emotional and behavioral measures (SCL-10); panel **c)** shows the performance of the model that excluded subjects (who did not have psychosis) with any ICD-10 F chapter diagnoses, and panel **d)** shows the performance of the model with this subset of subjects where we additionally adjusted CAPE-16 features for SCL-10.

**Table S9.** *Detailed results from the four CAPE-16 prediction models*

|  | **Regular model** | | | | | | | **Permutation testing** | | | | | | |
| --- | --- | --- | --- | --- | --- | --- | --- | --- | --- | --- | --- | --- | --- | --- |
| **Models** | **Sensitivity** | **Specificity** | **Accuracy** | **Avg. FP** | **Avg. TP** | **Avg. FN** | **Avg. TN** | **Sensitivity (*p*)** | **Specificity (*p*)** | **Accuracy (*p*)** | **Avg. FP** | **Avg. TP** | **Avg. FN** | **Avg. TN** |
| All subjects | 68.65 ± 5.21 | 82.39 ± 0.46 | 75.52 ± 2.61 | 2857 | 18 | 9 | 13362 | 37.69 ± 12.04 | 61.72 ± 4.88 | 49.71 ± 6.31 | 6209 | 10 | 17 | 10010 |
| All subjects,  SCL-10 regressed | 65.75 ± 4.90 | 83.10 ± 0.59 | 74.42 ± 2.37 | 2742 | 18 | 9 | 13477 | 37.11 ± 10.46 | 62.75 ± 2.92 | 49.93 ± 5.79 | 6042 | 10 | 17 | 10177 |
| Subject subset | 69.76 ± 4.00 | 83.85 ± 0.45 | 76.81 ± 1.98 | 2237 | 19 | 8 | 11613 | 38.85 ± 11.42 | 61.55 ± 5.27 | 50.20 ± 6.23 | 5326 | 11 | 16 | 8524 |
| Subject subset,  SCL-10 regressed | 66.51 ± 4.73 | 85.37 ± 0.48 | 75.94 ± 2.31 | 2026 | 18 | 9 | 11824 | 36.69 ± 11.63 | 63.61 ± 3.63 | 50.15 ± 6.78 | 5040 | 10 | 17 | 8810 |

*Note.* In exploratory analyses, we applied the RUSBoost algorithm to predict psychosis using CAPE-16. The analyses involved 48 features: Frequency items, distress items, and interaction features. Four models were run: Two with the complete sample and two with non-psychotic psychiatric diagnoses removed, each with and without current emotional symptoms (SCL-10) regressed in. Additionally, we conducted 100 permutation tests to assess if the prediction estimates were above chance level. SCL-10 = the Symptoms Checklist 10; Avg. = average; FP = false positives; TP = true positives; FN = false negatives; TN = true negatives.
